# Sleep Apnea and Hypertension Control among Hispanic/Latino Adults in the United States: Results from the Hispanic Community Health Study / Study of Latinos (HCHS/SOL)

**DOI:** 10.1101/2024.05.13.24307315

**Authors:** Cecilia Castro-Diehl, Amber Pirzada, Lisa de las Fuentes, Daniela Sotres-Alvarez, Carmen R. Isasi, Ramon A Durazo-Arvizu, Nour Makarem, Krista M. Perreira, Alberto R. Ramos, Sylvia Wassertheil-Smoller, Katie Stamatakis, Ariana M. Stickel, Susan Redline, Martha L. Daviglus

**Author notes:** **Corresponding Author:** Cecilia Castro-Diehl, MPH, DrPH.

## Abstract

**Objectives:** Hispanic/Latino adults have a high prevalence of uncontrolled hypertension predisposing them to CVD. We hypothesize that sleep apnea severity is associated with uncontrolled blood pressure (BP) and resistant hypertension in Hispanic/Latino adults.

**Methods:** This was a cross-sectional study of 2,849 Hispanic Community Health Study/Study of Latinos participants with hypertension (i.e., systolic BP ≥130 mm Hg, or diastolic BP ≥80 mm Hg or self-reported antihypertensive medication use) who were taking at least one class of antihypertensive medication. Participants were categorized as having *controlled (BP < 130/80 mmHg* among those on hypertension treatment)*, uncontrolled (BP* ≥ 130/80 mmHg using one or two classes of antihypertensive medications), or *resistant hypertension (BP* ≥ 130/80 mmHg while on ≥ 3 classes of antihypertensive medications or the use of ≥ 4 classes of antihypertensive medications regardless of BP control). Sleep apnea was classified based on the respiratory event index (REI; events/h) as *mild* (REI ≥ 5 and < 15), moderate-to-severe (REI ≥ 15), or *no sleep* apnea (REI < 5).

**Results:** In multinomial logistic regression, moderate-to-severe sleep apnea (vs. no sleep apnea) was associated with higher odds of resistant hypertension (Odds Ratio [OR], 2.15; 95% CI, 1.36-3.39 at 4% desaturation and OR 1.68; 95% CI, 1.05-2.67 at 3% desaturation). Neither mild nor moderate-to-severe sleep apnea was associated with uncontrolled hypertension.

**Conclusion:** Among diverse Hispanic/Latino persons, moderate-to-severe but not mild sleep apnea was associated with resistant hypertension. Identification and management of sleep apnea in this population may improve BP control and subsequently prevent adverse cardiovascular outcomes.

## Introduction

In the United States (US), racial and ethnic disparities in hypertension treatment and control persist and drive CVD inequities [1]. Although Hispanic/Latino adults have similar hypertension rates to non-Hispanic/Latino white adults per NHANES 2015-2018 data [2], they have a higher prevalence of uncontrolled hypertension, placing them at increased risk of future cardiovascular disease (CVD) [3]. Between 2008-18, the number of deaths attributable to high blood pressure (BP) increased by 106% in Hispanic/Latino adults [4]. Further, the Hispanic Community Health Study / Study of Latinos (HCHS/SOL), the largest cohort of Hispanic/Latino adults from diverse backgrounds in the US, has shown that the prevalence of hypertension varies substantially by Hispanic/Latino background, ranging from 17% among individuals of South American background to 34% among those of Dominican background [5]. In fact, Hispanic/Latino adults of Caribbean descent, a population with greater African ancestry genetic admixture, have similar hypertension prevalence and incidence rates to non-Hispanic/Latino Black adults, the group with the highest hypertension burden nationwide [4] [6].

Untreated sleep apnea is a known risk factor for hypertension [7, 8]. Mounting evidence suggests that sleep apnea may also be linked with poor BP control in those with treated hypertension [9]. Several studies have reported an association between sleep apnea and resistant hypertension, a particularly severe form of hypertension in which BP remains uncontrolled despite the use of at least three different classes of antihypertensive medications [10, 11]. However, previous studies are limited by modest sample sizes [10], study populations from clinical settings [12], and have focused on predominantly non-Hispanic Black and White adults [13]. To date, there are no population-based studies of sleep apnea severity in relation to resistant and uncontrolled hypertension in a population-based sample of Hispanic/Latino adults.

This study examined the association between overnight objectively recorded sleep apnea and uncontrolled or resistant hypertension among diverse Hispanic/Latino individuals with treated high blood pressure. We hypothesized that sleep apnea is associated with uncontrolled hypertension and that more severe sleep apnea would be associated with poorer BP control and greater odds of resistant hypertension.

## METHODS

### Study Sample

From 2008-2011, the HCHS/SOL enrolled 16,415 Hispanic/Latino men and women, aged 18–74 years from 4 communities in the United States (Bronx, NY; Chicago, IL; Miami, FL; San Diego, CA). Participants self-identified as Cuban, Puerto Rican, Dominican, Mexican, Central or South American, or other Hispanic/Latino origin. The study used a two-stage area probability sample of households with stratification by Hispanic/Latino concentration and proportion of high/low socio-economic status and over-sampling of individuals ages 45 years and older. Participants underwent a clinical examination and laboratory tests and completed questionnaires about their medical history and health behaviors. Participants were asked to bring all prescription and nonprescription medications taken in the past 4 weeks; medication names and dosages were noted on an inventory. Institutional review boards approved the study at all participating institutions, and all participants gave written informed consent.

The current study includes individuals with complete data on measures of sleep apnea and seated resting BP at Visit 1 (baseline exam) who took at least one class of hypertensive medication based on a review and inventory of medications by the study staff. Of the 16,415 individuals, 7,283 had hypertension (defined by the 2017 ACC/AHA BP guidelines [15], i.e., SBP ≥ 130 mmHg or DBP ≥ 80 mmHg or self-reported use of antihypertensive medications), and of these, 3,464 had treated hypertension (including self-reported antihypertensive medication use), and comprised our study population of interest. After excluding those who did not have objective data of sleep apnea or did not use at least one class of antihypertensive medication: angiotensin-converting enzyme inhibitors, angiotensin II receptor antagonists, beta-blockers, calcium channel blockers, or diuretics, 2,849 individuals were included in this analysis (**Supplementary** Figure 1).

### Ascertainment of Sleep Apnea Severity

Data on sleep apnea [14] was obtained using a home sleep apnea testing (HSAT) monitor (Apnea Risk Evaluation System [ARES], Unicorder, Advanced Brain Imaging, Carlsbad, CA). Using correlation analysis (r), others have shown excellent agreement with sleep apnea classification using this monitor and polysomnography (r=0.96 for in-laboratory ARES and r=0.88 for in-home ARES).[15] The monitor collected measures of overnight oxygen saturation, nasal airflow, snoring sounds, and head position and movement. Certified polysomnologists identified respiratory events, defined as a 50% or greater reduction in airflow lasting at least 10 seconds and associated with a ≥3% oxyhemoglobin desaturation after annotating artifacts and identifying probably sleep/wake epochs as described elsewhere. [16] Sleep apnea was assessed by using the respiratory event index (REI), which was calculated as the sum of the number of apneas and hypopneas associated with 3% (REI3%) or 4% (REI4%) oxygen desaturation divided by the total sleep time [events/h]). Presence/severity of sleep apnea was categorized as no sleep apnea (REI < 5 events/h; reference group), *mild* (REI ≥ 5 and < 15), and *moderate-to-*severe (REI ≥ 15). We considered sleep apnea associated with 4% (REI4%) and with ≥3% desaturation (REI3%) oxyhemoglobin desaturation.

### Measurement of Blood Pressure and Definition of Uncontrolled and Resistant Hypertension

Seated BP was measured using a standard protocol after 5 minutes of rest. Three measurements were obtained one minute apart, and the average of the three measures was used. *Hypertension treatment* was defined as the use of at least one antihypertensive medication during the 4 weeks before the clinic examination, based on an inventory of medications brought by the participant. Hypertension was defined based on the 2017 American College of Cardiology/American Heart Association (ACC/AHA) guidelines. [17] *Controlled hypertension* was defined as having SBP < 130 and DBP < 80 mmHg among those on hypertension treatment. *Uncontrolled hypertension* was defined as having SBP ≥ 130 or DBP ≥ 80 using one or two classes of antihypertensive medications (angiotensin-converting enzyme inhibitors, angiotensin II receptor antagonists, beta-blockers, calcium channel blockers, or diuretics). *Resistant hypertension* was defined as having SBP ≥ 130 or DBP ≥ 80 while on ≥ 3 classes of antihypertensive medications, with one class being a diuretic or the use of ≥ 4 classes of antihypertensive medications regardless of BP control.

In an extended analysis, we also defined *hypertension* based on the Seventh Report of the Joint National Committee (JNC 7) of high blood pressure guidelines [18] as average SBP ≥ 140 mm Hg or average DBP ≥ 90 mm Hg or use of antihypertensive medication. Only participants who were taking antihypertensive medications in the 4 weeks prior to their clinic visit based on medication inventory were included (n = 2884). *Controlled hypertension* was defined as SBP < 140 mmHg and DBP < 90 mmHg among those taking hypertension medication. *Uncontrolled hypertension* was defined as SBP ≥ 140 or DBP ≥ 90 and the use of one or two classes of antihypertensive medications. *Resistant hypertension* was defined as SBP ≥ 140 or DBP ≥ 90 while on ≥ 3 classes of antihypertensive medications, with one class being a diuretic or the use of ≥ 4 classes of antihypertensive medications regardless of BP control.

### Assessment of Covariates

Participants’ age, sex, Hispanic/Latino background, site, health insurance status (yes, no), and education were ascertained by self-report. Education was categorized as less than high school, high school, or general equivalency diploma and greater than high school or GED (bachelor or higher). Smoking status was categorized as current, former, or never. We also included clinical parameters: Body mass index (BMI, kg/m^2^) was calculated as measured weight in kilograms divided by the square of height in meters, and obesity was defined as a body mass index ≥ 30 kg/m^2^. *Diabetes* was defined based on the American Diabetes Association criteria [19], i.e., fasting glucose ≥ 126 mg/dL if fasting time > 8h or fasting glucose ≥ 200 mg/dL if fasting time ≤ 8h or post-OGTT test≥ 200 mg/dL or A1C≥ 6.5% or self-report of physician-diagnosed diabetes. *Prediabetes* was defined as having fasting glucose 100-125 mg/dL (if fasting time >8h) or post-OGTT test 140-199 mg/dL or 5.7%≤A1C<6.5%. Insomnia symptoms were derived from the Women’s Health Initiative Insomnia Rating Scale (WHIIRS). [20]

### Statistical Analysis

All analyses accounted for HCHS/SOL complex survey design (stratification, clustering, and sampling weights) using survey procedures in SAS 9.4. Among individuals with hypertension and receiving antihypertensive medications, we described baseline demographic, socioeconomic, clinical, and behavioral characteristics within each sleep apnea category. T-test and Chi-squared tests or Wilcoxon rank sum or Kruskal Wallis tests were conducted to evaluate differences in the continuous and categorical variables distribution across the sleep apnea groups.

Multinomial logistic regression was used to examine the association of sleep apnea (mild and moderate-to-severe sleep apnea vs. none [referent category]) with uncontrolled hypertension or resistant hypertension vs. controlled [referent category]) among persons with hypertension and receiving antihypertensive medication. The odds ratio (OR) and 95% confidence interval (OR and 95% CI) were computed. We used a hierarchical approach to adjust our models. Model 0 was an unadjusted model. Model 1 was adjusted for age and site. Model 2 included covariates in model 1 plus sex, Hispanic/Latino background, education, health insurance, and smoking status. Model 3 was adjusted for covariates included in model 2 plus BMI.

## RESULTS

### Sleep Apnea and Hypertension Prevalence

**Table 1** describes baseline characteristics of 2849 Hispanic/Latino individuals with treated hypertension by sleep apnea severity. About 23% of individuals had mild sleep apnea, 14% had moderate-to-severe sleep apnea, and 63% did not have sleep apnea as defined by REI <5.

**Table 1.**
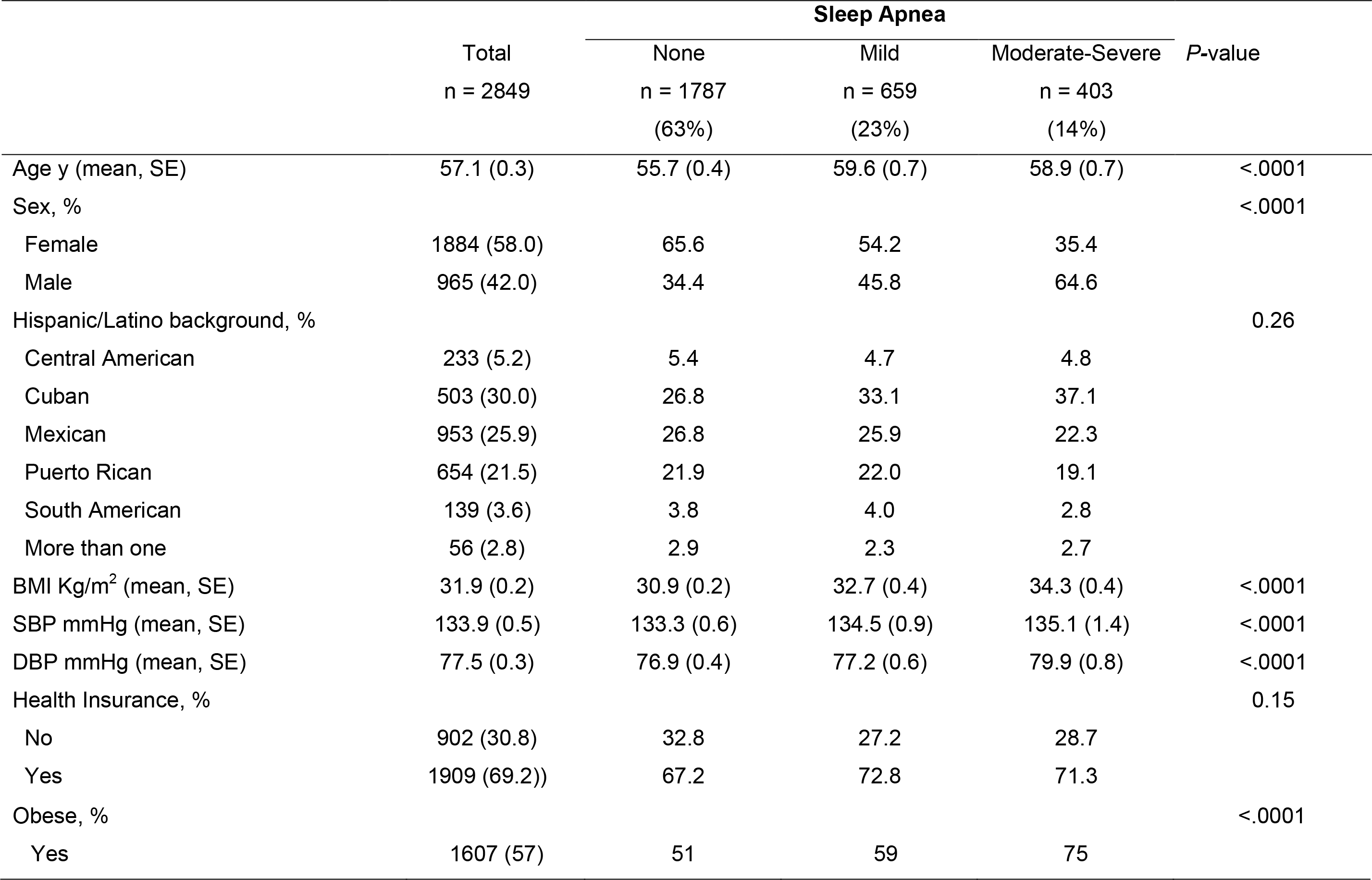
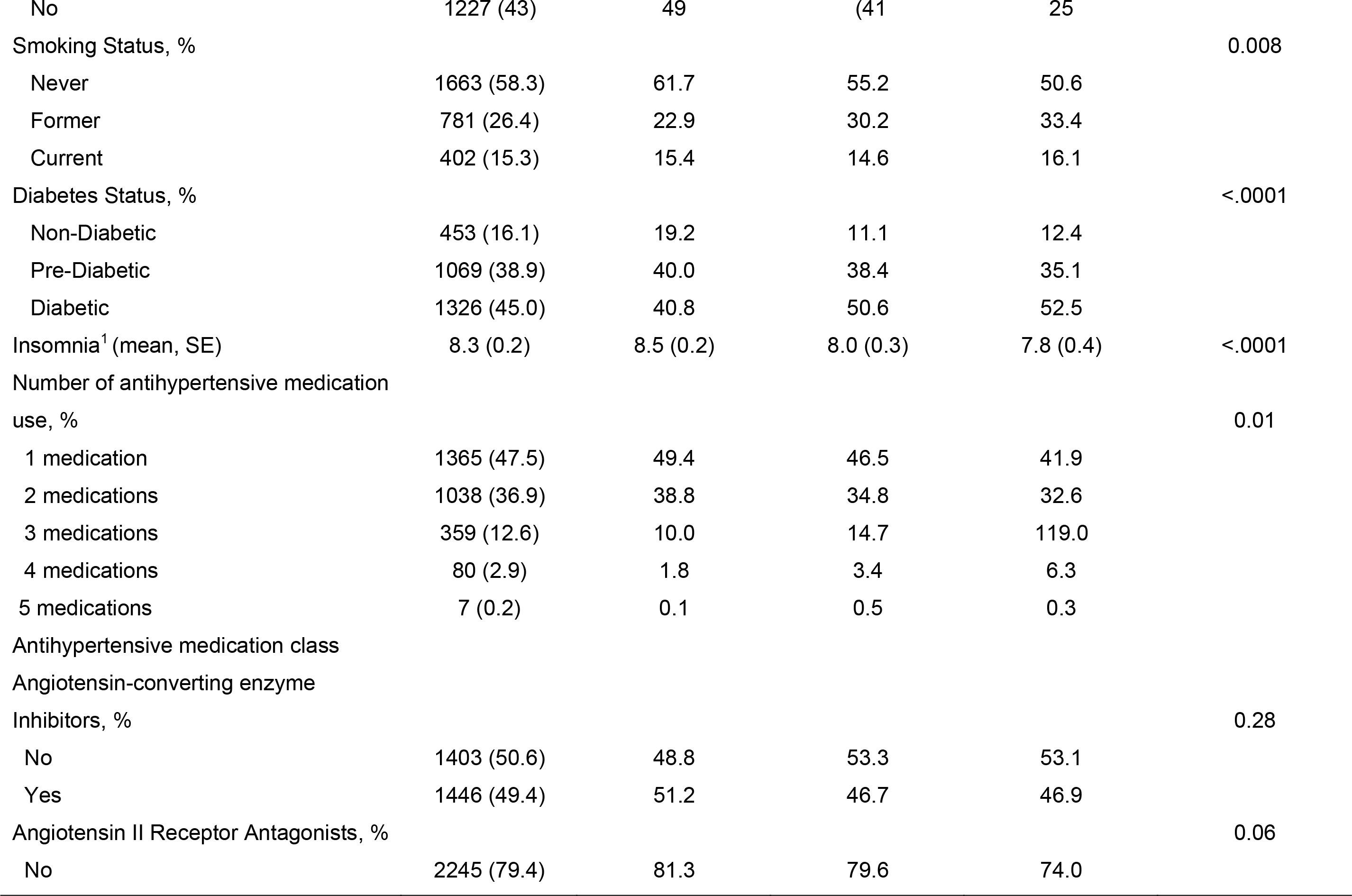
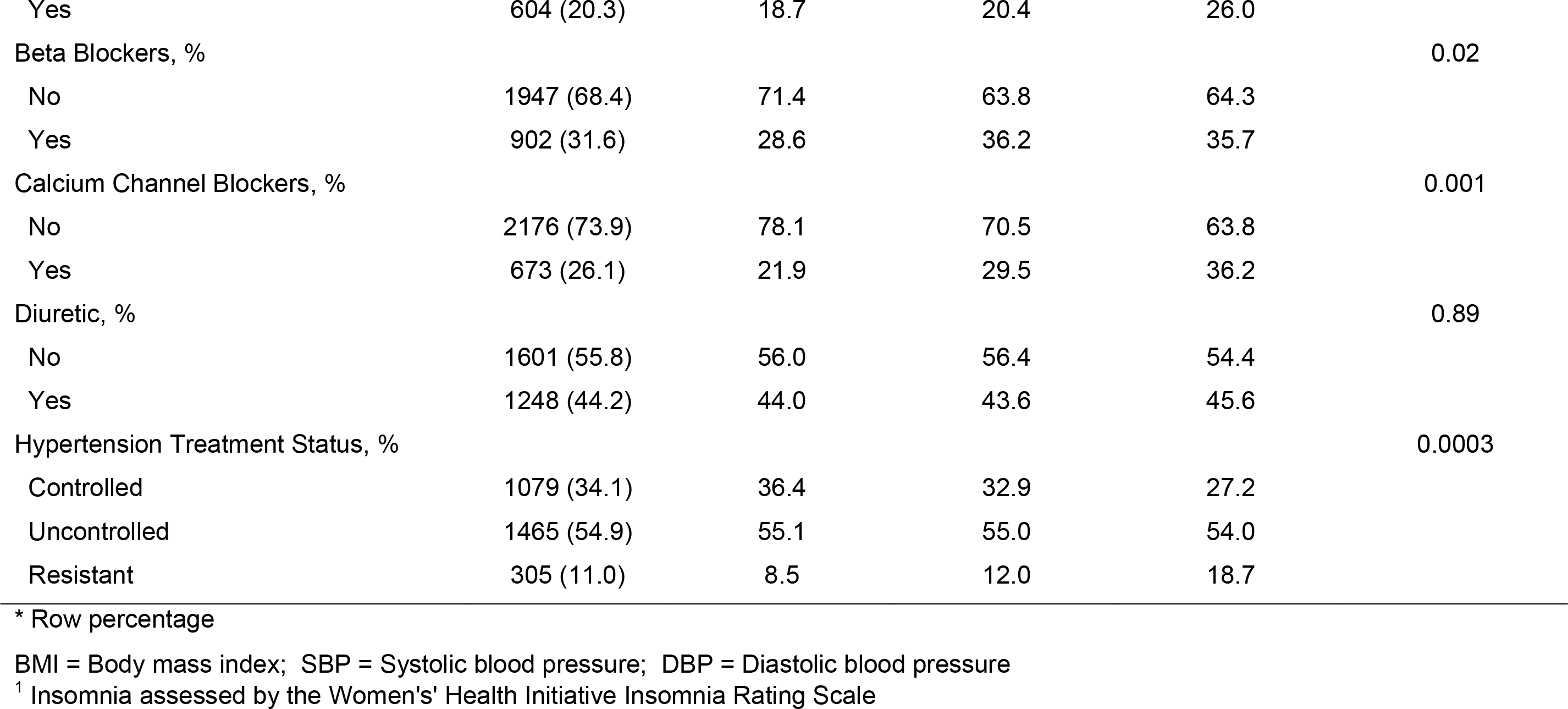
Sociodemographic, Behavior and Health Characteristics, by Sleep Apnea Severity (n=2959), HCHS/SOL (2008-2011)

Sleep apnea prevalence was higher in older vs. younger individuals. Men were more likely than women to have greater sleep apnea severity. Similarly, former and current smokers were more likely than never smokers to have moderate-to-severe sleep apnea. Sleep apnea severity increased with higher BMI. There were no differences in sleep apnea severity by Hispanic/Latino background or health insurance. A very small percentage of the study individuals (∼1%) reported a physician diagnosis of sleep apnea (**data not shown**).

Overall, our study population included 34%, 55%, and 11% individuals with controlled, uncontrolled, and resistant hypertension, respectively. **Figure 1** shows the prevalence of controlled, uncontrolled, and resistant hypertension by sex and age group.

**Figure 1.**
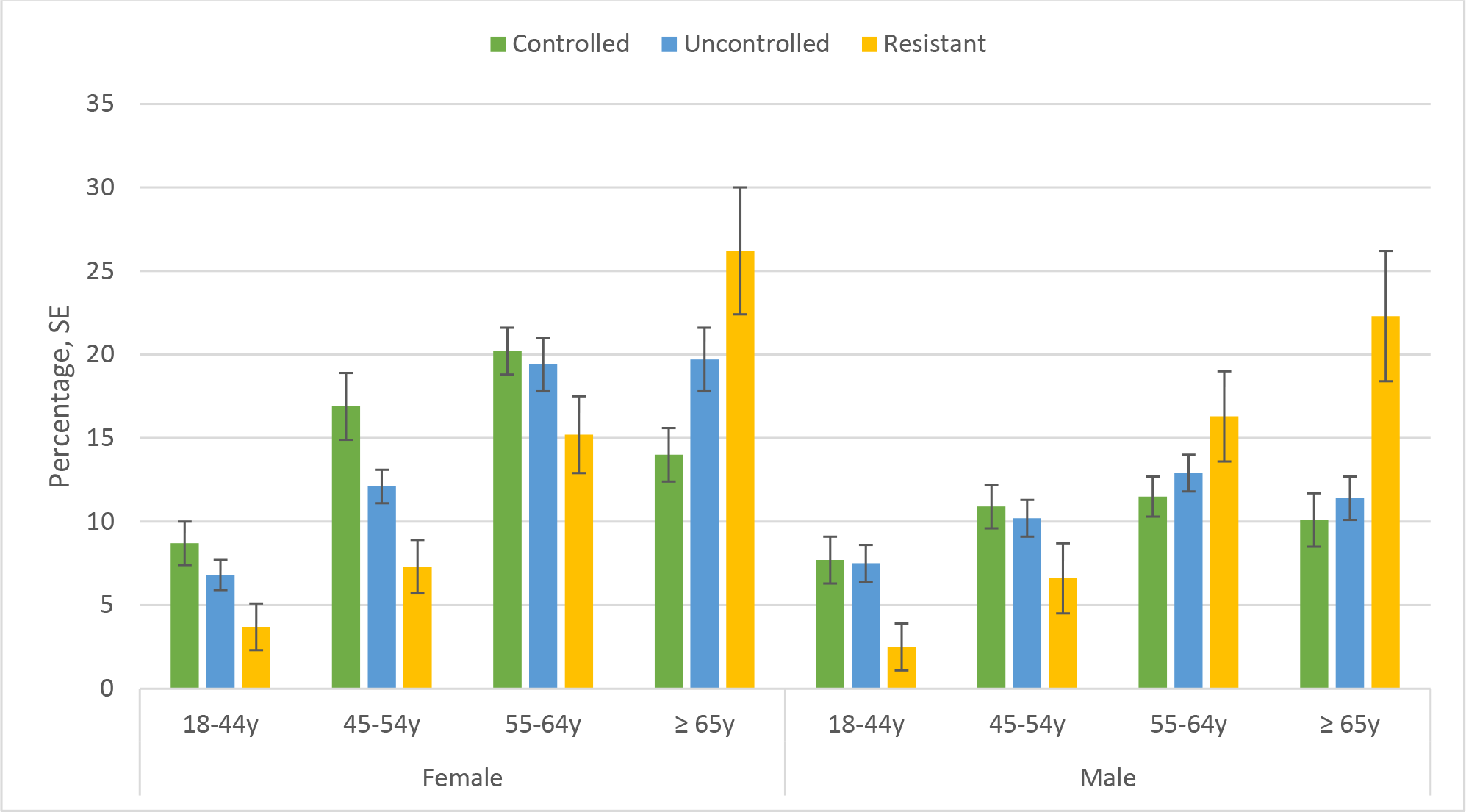
Prevalence of Controlled, Uncontrolled and Resistant Hypertension among Individuals with Treated Hypertension, HCHS/SOL (2008-2011).

### Association of Sleep Apnea Severity with Uncontrolled and Resistant Hypertension using the definition of Hypertension by the 2017 ACC/AHA

In multinomial logistic regression analyses, moderate-to-severe, but not mild, sleep apnea (based on respiratory events requiring a minimum of 4% desaturation, REI4%) was associated with higher odds of resistant hypertension relative to no sleep apnea (OR, 2.15; 95% CI, 1.36- 3.39; p=0.001) (**Table 2**) after full covariates adjustment. Effect estimates were attenuated but remained statistically significant when sleep apnea was defined based on respiratory events requiring a minimum of 3% desaturation. Moderate-to-severe sleep apnea (based on REI3% desaturation) was associated with higher odds of resistant hypertension relative to no sleep apnea (OR, 1.68; 95% CI, 1.05-2.67; p = 0.03) (**Table 3**). Neither mild nor moderate-to-severe sleep apnea was associated with uncontrolled hypertension in any models with apnea based on 4% or 3% desaturation events.

**Table 2.**
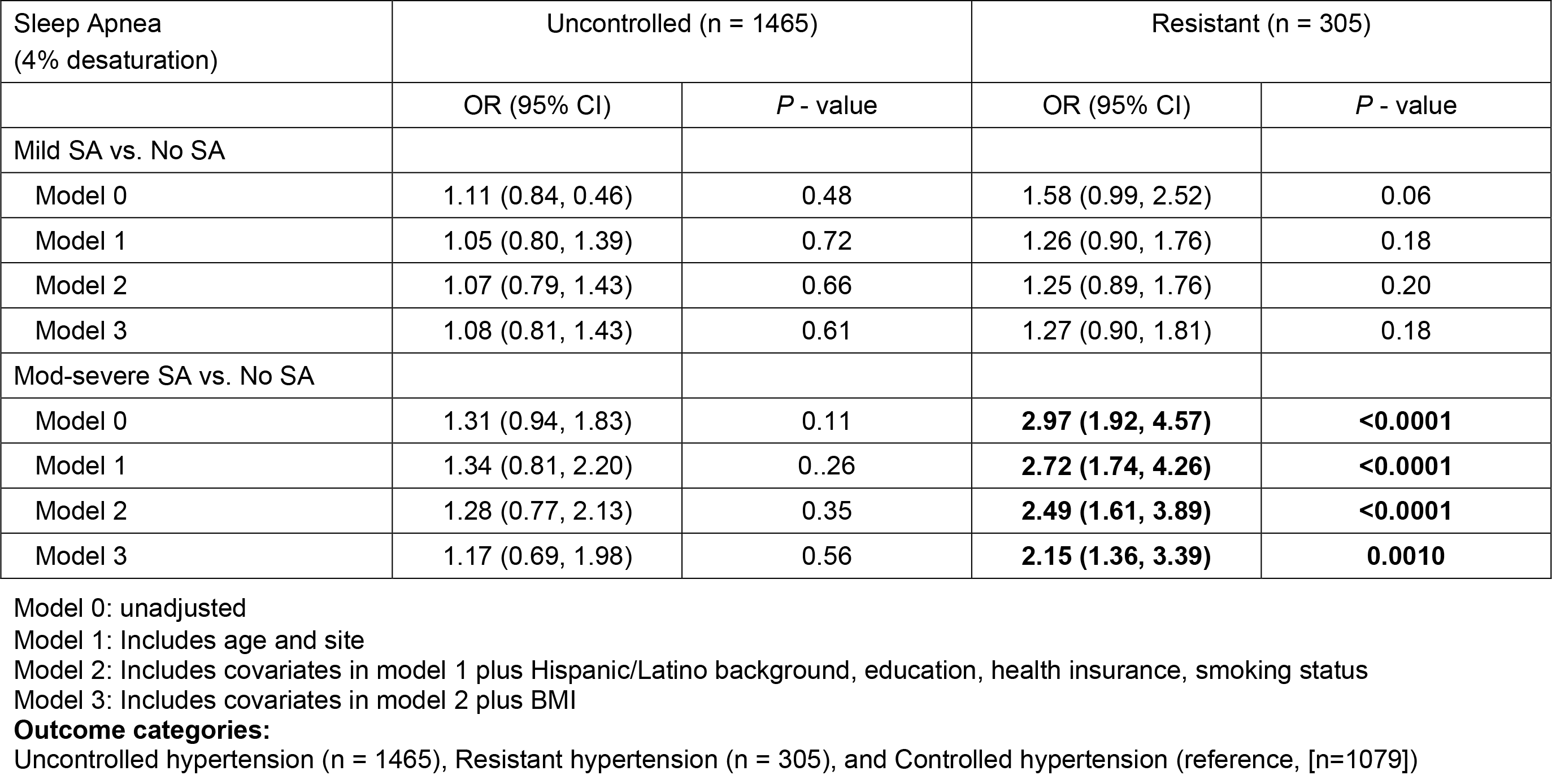
Odds Ratios (OR [95% Confidence Intervals, CI])) for Associations of Mild and Moderate-to-Severe Sleep Apnea defined by 4% desaturation with Uncontrolled or Resistant Hypertension by 2017 ACC/AHA Definition vs. Controlled Hypertension, HCHS/SOL (2008-2011).

| Sleep Apnea<br>(4% desaturation) | Uncontrolled (n = 1465) |  | Resistant (n = 305) |  |
| --- | --- | --- | --- | --- |
|  | OR (95% CI) | P - value | OR (95% CI) | P - value |
| Mild SA vs. No SA |  |  |  |  |
| Model 0 | 1.11 (0.84, 0.46) | 0.48 | 1.58 (0.99, 2.52) | 0.06 |
| Model 1 | 1.05 (0.80, 1.39) | 0.72 | 1.26 (0.90, 1.76) | 0.18 |
| Model 2 | 1.07 (0.79, 1.43) | 0.66 | 1.25 (0.89, 1.76) | 0.20 |
| Model 3 | 1.08 (0.81, 1.43) | 0.61 | 1.27 (0.90, 1.81) | 0.18 |
| Mod-severe SA vs. No SA |  |  |  |  |
| Model 0 | 1.31 (0.94, 1.83) | 0.11 | **2.97 (1.92, 4.57)** | **<0.0001** |
| Model 1 | 1.34 (0.81, 2.20) | 0.26 | **2.72 (1.74, 4.26)** | **<0.0001** |
| Model 2 | 1.28 (0.77, 2.13) | 0.35 | **2.49 (1.61, 3.89)** | **<0.0001** |
| Model 3 | 1.17 (0.69, 1.98) | 0.56 | **2.15 (1.36, 3.39)** | **0.0010** |

Model 0: unadjusted
Model 1: Includes age and site
Model 2: Includes covariates in model 1 plus Hispanic/Latino background, education, health insurance, smoking status
Model 3: Includes covariates in model 2 plus BMI

**Table 3.** Odds Ratios (OR [95% Confidence Intervals, CI]) for Associations of Mild and Moderate-to-Severe Sleep Apnea defined by 3% desaturation with Uncontrolled or Resistant Hypertension by 2017 ACC/AHA Definition vs. Controlled Hypertension, HCHS/SOL (2008-2011).

| Sleep Apnea<br>(3% desaturation) | Uncontrolled (n = 1465) |  | Resistant (n = 305) |  |
| --- | --- | --- | --- | --- |
|  | OR (95% CI) | *P* - value | OR (95% CI) | *P* - value |
| Mild SA vs. No SA |  |  |  |  |
| Model 0 | 1.07 (0.80, 1.44) | 0.63 | **1.36 (1.002, 1.85)** | **0.04** |
| Model 1 | 1.01 (0.75, 1.36) | 0.96 | 1.27 (0.94, 1.73) | 0.12 |
| Model 2 | 0.99 (0.73, 1.34) | 0.94 | 1.28 (0.93, 1.74) | 0.13 |
| Model 3 | 0.93 (0.73, 1.33) | 0.91 | 1.28 (0.94, 1.77) | 0.12 |
| Mod-severe SA vs. No SA |  |  |  |  |
| Model 0 | 1.25 (0.78, 1.99) | 0.35 | **2.51 (1.63, 3.86)** | **<0.0001** |
| Model 1 | 1.05 (0.65, 1.71) | 0.84 | **2.11 (1.35, 3.30)** | **0.001** |
| Model 2 | 1.05 (0.64, 1.72) | 0.85 | **1.99 (1.28, 3.09)** | **0.002** |
| Model 3 | 0.96 (0.58, 1.60) | 0.89 | **1.68 (1.05, 2.67)** | **0.03** |

Model 0: unadjusted
Model 1: Includes age and site
Model 2: Includes covariates in model 1 plus Hispanic/Latino background, education, health insurance, smoking status
Model 3: Includes covariates in model 2 plus BMI

### Association of Sleep Apnea Severity with Uncontrolled and Resistant Hypertension using the definition of Hypertension by the JNC 7 Guidelines

When we examined these associations using hypertension defined per the JNC 7 guidelines, moderate-to-severe, but not mild, sleep apnea (based on REI4% desaturation) was associated with higher odds of resistant hypertension relative to no sleep apnea (OR, 1.94; 95% CI, 1.18- 3.17; p=0.01) (**Table S1**) after full covariates adjustment. Sleep apnea based on REI3% desaturation was not associated with resistant or uncontrolled hypertension **(Table S2)**.

## DISCUSSION

In the current study of Hispanic/Latino adults of diverse backgrounds with treated hypertension, the prevalence of mild and moderate-to-severe sleep apnea was 23% and 14%, respectively, which is within the range of the general adult population [21]. Uncontrolled and resistant hypertension among individuals with treated hypertension was present in 55% and 11% of the study population, respectively. Moderate-to-severe sleep apnea was associated with up to two-fold higher odds of resistant hypertension but not uncontrolled hypertension. These results underscore the importance of treating sleep apnea for prevention of resistant hypertension and its downstream influence on future CVD risk.

The population-based Wisconsin Study conducted more than two decades ago in a non-Hispanic White population showed a dose-response relationship between untreated sleep-disordered breathing and hypertension independent of age, sex, and adiposity. [22]. Since then, other epidemiological studies in white populations have replicated these findings [23–25].

Previous studies have also reported that the prevalence of sleep apnea in persons with resistant hypertension was high, ranging from 72% [26] to more than 80% [27–30], and suggested that more severe sleep apnea was associated with a lower likelihood of BP control even with greater antihypertensive medication use [10, 23, 25]. However, these studies had relatively small sample sizes (n=41 to 422 participants) [30], were conducted in predominantly non-Hispanic/Latino Whites in specific settings (clinical vs. epidemiological studies), and used differing criteria to define sleep apnea severity, which limits the comparability of findings across studies. In addition to the use of nonstandard criteria to define sleep apnea, the definition of hypertension, uncontrolled, and resistant hypertension has also varied across studies and has changed with time.

Our findings are consistent with a retrospective cohort study of a racially, ethnically, and socioeconomically diverse population from California [31, 32] in showing a higher rate of sleep apnea among individuals with resistant hypertension than among those without resistant hypertension (9.6% vs. 6.8%) and greater odds of sleep apnea with resistant hypertension vs. non-resistant hypertension (OR 1.16 [95% CI 1.12, 1.19]). However, that study did not specifically focus on Hispanic/Latino populations, and our findings reported herein highlight that these associations may be stronger in these population groups than previously reported. In fact, we report a similar effect size to that observed among Black adults in the Jackson Heart Study [13]. Although the prevalence of resistant hypertension (15%) was higher in the Jackson Heart study than the current study (11%) [17] those with moderate-to-severe sleep apnea also had two times higher odds of resistant hypertension (OR, 2.00; 95% CI,1.14-3.67) compared to those without sleep apnea [13].

Mechanisms that may explain the association between sleep apnea and hypertension include intermittent hypoxia [33], frequent intermittent sympathetic stimulation, a blunted baroreflex, oxidative stress, metabolic abnormalities, arterial stiffness, endothelial dysfunction [34], which also may explain the association between sleep apnea and poorly controlled hypertension. Non-dipping BP is particularly common in sleep apnea and often coexists in patients with resistant hypertension. Indeed, there is a significant association between resistant hypertension, non-dipping BP, and endothelial dysfunction [34]. Aldosterone excess is common in persons with resistant hypertension and contributes to the severity of sleep apnea. Aldosterone will induce fluid retention, leading to parapharyngeal edema and increase upper airway resistance [35, 36].

Obesity is linked with more severe hypertension and lack of control of hypertension besides an increasing number of antihypertensive medications [37]. This relation may be explained by impaired sodium excretion and activation of the renin-angiotensin-aldosterone system [37] [38].

Obesity is also a recognized risk factor for sleep apnea [33]. In the current study, the association between sleep apnea and resistant hypertension was attenuated but remained significant with adjustment for BMI.

The null association between sleep apnea and uncontrolled hypertension may be explained by the definition used for uncontrolled hypertension, which presents a challenge in its interpretation. Some persons with uncontrolled hypertension may not have been identified, because they remained undiagnosed and untreated [39]. Persons with resistant hypertension, which is determined by more strict definitions, are a subset of a larger, less well-defined group of persons with uncontrolled hypertension that may additionally include those with poor medication adherence and/or poor medical management [40]. In addition, both uncontrolled and resistant hypertension could also be affected by masked hypertension, the effect of having normal BP in-office visits but high BP at home. Our study, similar to other studies [12, 13], did not find an association between sleep apnea and uncontrolled hypertension.

Nonetheless, our study has several potential limitations. First, these analyses were cross-sectional, which limited our ability to establish temporality and infer causality. Second, the sleep study device used allowed us to define sleep-disordered breathing but not to differentiate obstructive sleep apnea from central sleep apnea; however, in this relatively young, community-based cohort the prevalence of central sleep apnea is likely very low. Third, other studies have used ambulatory BP devices to define or confirm resistant hypertension, whereas our study used three measurements of office BP taking during a single examination visit, which is more prone to the effects of white-coat or masked hypertension. Because of our sample size, we had limited power to compare differences in associations across Hispanic/Latino groups. Finally, we defined resistant hypertension based on the 2017 ACC/AHA guidelines in a population who attended their study examination when the JNC 7 guidelines were still in place. This could inflate our prevalence estimates of resistant hypertension since physicians would be trying to target BP control at a cut-off of 140/90 mmHg instead of a lower cut/off (130/80 mmHg).

However, our study also has several strengths. First, this is the first study of the association of sleep apnea and resistant hypertension in a population-based cohort of diverse Hispanic/Latino adults in the US. Second, we used home sleep testing to assess sleep-disordered breathing objectively. Third, participants brought the bottles of medications they were using during the four weeks prior to their visit for scanning and inclusion in the participant record. Fourth, HCHS/SOL collected comprehensive measures of lifestyle, social, and medical factors that allowed us to adjust our analyses for several confounders. Finally, we examined the relationship between sleep apnea and uncontrolled elevated BP using both 130/80 and 140/90 mmHg thresholds, which enhances comparability of our results to previous and more recent studies.

## Conclusions

In conclusion, among diverse Hispanic/Latino adults, moderate-to-severe sleep apnea compared to no sleep apnea was associated with prevalent resistant hypertension, defined as uncontrolled hypertension despite aggressive medication use. These findings suggest that moderate-to-severe sleep apnea may play a role in poor hypertension control among persons receiving intensive antihypertensive treatment. Our findings warrant confirmation prospectively to determine the influence of sleep apnea on the incidence of uncontrolled and resistant hypertension in the US Hispanic/Latino population. Given the increased cardiovascular risks posed by sleep apnea and considering our study’s findings on the relation with resistant hypertension, diagnosis, and management of sleep apnea may be an important target for enhanced BP control and improving cardiovascular outcomes in this population.

## Data Availability

The Hispanic Community Health Study / Study of Latinos (HCHS/SOL) data used to support the findings of this study are available at the HCHS/SOL Coordinating Center at request after following procedures. HCHS/SOL Website https://sites.cscc.unc.edu/hchs/

## Acknowledgments

The authors thank the staff and participants of HCHS/SOL for their important contributions. A complete list of staff and investigators has been provided by Sorlie P., et al. in Ann Epidemiol. 2010 Aug;20: 642-649 and is also available on the study website http://www.cscc.unc.edu/hchs/

